# Evaluation of The Pink Luminous Breast LED-Based Technology Device as a Screening Tool for the Early Detection of Breast Abnormalities

**DOI:** 10.1101/2021.03.23.21253758

**Authors:** Fernando Ocasio-Villa, Luisa Morales-Torres, Norma Velez-Medina, Juan C. Orengo, Edu B. Suarez Martinez

## Abstract

Breast cancer is the leading cause of sex-specific female cancer death in the United States. Detection at earlier stages contributes to decrease the mortality rate. The mammography is considered the gold standard for breast cancer screening with an estimate sensitivity of 86.9% and a specificity 88.9%. However, these values are negatively affected by the breast, which is consider a risk factor for developing breast cancer. Herein, we validate the novel LED-based FDA Class I medical device Pink Luminous Breast (PLB) by the comparison of two breast screening imaging-based tests using a double blinded approach. The PLB works by emitting a LED red light with a harmless spectrum of 640-800 nanometers, the trans-illuminated breast tissue allows the observation of abnormalities represented by darker or shadowing areas. In this study, we evaluated the sensitivity and specificity of the PLB device as a screening tool for the early detection of breast abnormalities when compared with the mammography as the gold standard. Our results showed that PLB device has a high sensitivity (89.6%) and specificity (96.4%) for detecting breast abnormalities comparable to the adjusted mammography values: 86.3% and 68.9% respectively. Importantly, the percentage of positive dense tissue findings from a total of 340 events was 266 (78.2%) using PLB vs. 248 (72.9%) detected by the mammography. A 100% of the participants responded in a survey that they feel comfortable using the device and visualizing their breast without feeling pain or discomfort during the examination. The PLB positive validation vs the mammography brings the potential to be recommended for routinely breast screening at non-clinical settings. The PLB provides a rapid, non-invasive, portable, and easy-to-use tool for breast screening that can complement the home-based BSE technique or the CBE. In addition, the PLB can be conveniently used for screening breasts with surgical implants. PLB provides an accessible and painless breast cancer screening tool. The use of this device is not intended to replace the mammography as the gold standard for breast screening but rather to use it as an adjunct or complement tool as part of more efficient earlier detection strategies and contribute to decrease this health disparity.

## INTRODUCTION

Breast cancer is the number one female sex-specific cause of cancer death in the United States (American Cancer Society (ACS), 2021; Centers for Disease Control and Prevention (CDC), 2017). Despite therapeutic advances in the field, the medical and clinical community agree that early detection is the best approach to decrease the mortality rate of this disease (Welch, et al., 2016; World Health Organization, 2021; Duffy, et al., 2020; Tabar, et al., 2021). The objective of breast cancer screening is to detect the disease at a pre-clinical stage in asymptomatic patients to decrease the mortality rate and increase prognostic survival curves, avoiding extreme and expensive interventions with a negative effect in the patient’s quality of life. In the clinical practice, the mammography is the primary imaging-based test and considered as the gold standard screening test for identifying breast abnormalities, including suspicious lesions for breast cancer. As a public health-oriented screening test, the mammography is intended to be cost-effective and evidence-based accessible to the population. Previous studies identified socio-economic status, ethnicity, and health insurance coverage as major variables for the lack of primary screening and following up visits (Banks, et al., 2004; Tabar, et al., 2021). There are also cultural factors and misinformation within women regarding the importance, significance, and positive outcomes if they undergo routine examinations for detecting breast abnormalities at earlier stages such as increasing life expectancy and survival curves (Khan & Chollet, 2021). Although the clinical breast examination (CBE) by palpation combined with the mammography its sensitivity increased a 4% (Kuhl, et al., 2005; Banks, et al., 2004) it is not approved by most relevant clinical groups guidelines for breast screening U.S. Preventive Services Task Force (USPSTF), American Academy of Family Physicians (AAFP), ACS). However, the breast self-examination (BSE) is recommended for encouraging breast self-awareness by American College of Obstetricians and Gynecologists (ACOG), ACS, and National Comprehensive Cancer Network (NCCN). Interestingly, the scientific literature suggests that the mammography with a complementary secondary or third image-based device testing increases its sensitivity (Berg, et al., 2010; Buchberger, et al., 2018) and some concern about a decrease of the specificity with the Magnetic Resonance Imaging (MRI) (Kuhl, et al., 2005; Saslow, et al., 2007; Berg, et al., 2012; Vreeman, et al., 2018), none of other available imaging tools are included at the breast cancer screening guidelines for an average cancer risk person including insufficient evidence for dense breast outcomes (USPSTF, AAFP, ACS) and neither for women considered at high risk USPSTF, AAFP.

There are many studies comparing the sensitivity and specificity of the mammography with other imaging-based screening techniques such as digital mammography, ultrasonography, Tomosynthesis (3D), MRI, and even positron emission tomography (PET) scan (Berg, et al., 2006; Zhang, et al., 2014; Saslow, et al., 2007; Lee, et al., 2019; Mehnati & Tirtash, 2015;Myers, et al., 2016; Tagliafico, et al., 2018; Ezratty, et al., 2020). Together, these studies concurred that current imaging tests other than the mammography despite providing higher sensitivity, specificity for breast screening, require highly regulated settings and other costs-related expenses. These include contrasting solutions, prior patient preparations, and more qualified personnel among other. For instance, these other imaging-based tests provide a clinical added value as supplemental screening after the abnormal mammography results, for populations classified as having higher risk determined by a breast cancer assessment model, or when another underlying disease is clinically suspected (Sprague, et al., 2014; Khan & Chollet, 2021).

As the gold-standard screening test, abnormal mammography observations should generate awareness to promote an informed state of mind about the potential diagnosis and prognosis for the tested person. More importantly, abnormal results are intended to alert physicians about the risks representing these outcomes such as potential malignancies and ought to follow the recommended guidelines for patient standard of care on these cases (USPSTF, AAFP, and ACS). Concerningly, Ezratty, (2020) reported a three-years retrospective, observational cohort study of women living in New York USA reported non-Hispanics blacks and Hispanic women are less likely to receive a supplemental imaging-based testing follow up order, when compared to non-Hispanic white women. Her group also found that generalist physicians were less likely to appoint follow up visits and order supplemental imaging-based test, to these groups even when presenting suspicious or inconclusive mammography results, contrasting to specialty physicians (Ezratty, et al., 2020).

Although the mammography seems convenient as a breast abnormality screening tool (the benefits super pass the harms), this non-invasive test requires X-Rays exposure, regulated facilities, and trained personnel. It is also well-documented that one of the main reasons discouraging women to perform their scheduled mammography is the discomforting and pain experience during the testing (Nelson, et al., 2016). More importantly, it is questionable if the mammography is the best screening tool for women with dense or highly dense breast tissue, which is consider a breast cancer risk factor by the cancer assessment models (Berg, 2016; Feer, 2015. The former brings concerns to the public health and clinical community if there is an underestimation or misdiagnosed (increase false negative results) cases in women from 40-74 years of age, especially when those age ranges represent a 50% of dense breast tissue findings and dense breast is considered a risk factor for breast cancer (Sprague, et al., 2014; Melnikow, et al., 2016; Haas & Kaplan, 2015). Four instance, new technologies with similar or highest sensitivity and specificity capable to overcome cost expenses, cultural and religious believes, complaining about discomfort, and the potential false negative results on dense breast tissue is highly justified.

As a potential solution to mammography limitations other light-based devices have been tested since the 90’s but they showed low sensitivity and specificity (Alveryd, et al., 1990; Labib, et al., 2013; Shiryazdi, et al., 2015). Herein, we introduce the novel LED based and registered FDA class I medical device: The Pink Luminous Breast (PLB). The PLB works by emitting a LED red light with a harmless spectrum of 640-800 nanometers that is safely absorbed by hemoglobin, allowing the transillumination through the breast tissue and observations of abnormalities represented by darker or shadowing areas. In this study, we evaluated the sensitivity and specificity of The PLB device as a screening tool for the early detection of breast abnormalities and including cancer suspicious lesions when compared with the mammography as the gold standard.

## MATERIALS AND METHODS

A comparison of two diagnostic devices using a double blinded approach. The study comprised of 170 Puerto Rican women participants, randomly recruited from the general population *via* electronic communications including social media advertising, and personal communications responding to flyers displayed at physicians’ offices, local clothing stores, and drug stores. The study was conducted from June 2020 to December 2020 at Centro de Desarrollo de Investigaciones en Ciencias y Tecnología de Ponce, C.D. (CDICTP), Ponce, Puerto Rico. This study was approved by the Ponce Health Sciences University Institutional Review Board, Ponce, Puerto Rico as protocol #1911024753. In addition, we implemented preventive measurements for COVID-19 as stipulated by the Puerto Rico Governor’s Executive Order 2020-018 effective on May 1, 2020. The inclusion criteria of this study consisted of: Puerto Rican females over 40 years old with printed mammography results dated from March 2019 to December 2020 as certified by a radiologist. We used the following as exclusion criteria: persons receiving treatment of chemotherapy or radiotherapy, pregnant women, open wounds, infections, lacerations, scratches on the breast or nipple area, breastfeeding, hormonal treatment for fertility, history of autoimmune diseases, and a total or partial mastectomy, unilateral or bilateral.

Upon arrival, the CDICTP Scientific Coordinator (SC) welcomed and escorted the participants to the study office. All participants signed the IRB approved consent form prior to initiate the corresponding assessments. The SC requested each participant the printed mammography results to verify if the dates were valid as described at the inclusion criteria section. Then, the SC copied (Hewlett Packard Pro 477; Boelingen, Germany) the results and erase the name using a white eraser tape. The SC copied the results without the name, wrote a coding number to the last copy as it appeared in an envelope previously coded, placed the coded copy inside the matching coded envelop, sealed it, and stored it in a locked file cabinet with limited access. This last step is required for confidentiality purposes but in our case, we also ensured to minimized or avoid *bias* by the time of carrying out the blind analyses. We returned the original and the copy with white eraser tape to the participant. On that same line, we instructed the participants not to comment or discuss their mammography reports with any member of the research staff or other participants waiting for their appointment.

Then, the participants fulfilled a questionnaire and next were escorted by the SC to the examination room. Once in the examination room, the participant removed the upper side clothing and wore an examination paper gown open at the front. The Study Specialist (SS) presented a 3-minute instructional video showing the correct use of the PLB device. Following the video and prior to start the examination, the SS described to the participants all the steps to be performed *in-site* and informed the possibility of feeling discomfort such as cold to the touch. The non-invasive PLB device was properly disinfected before and after examination by using an antibacterial solution and 90% ethyl alcohol. The participant stood in front of a mirror at a distance of 2-feet with the room lights turned off. The SS first examined the right breast at three specific areas using the clock as reference for reporting in the following specific order: 6-o’clock, 3-o’clock, and 9-o’clock. The examination areas were documented by a photograph with a digital camera and saving the files under the corresponding code number then transferred as a digital file to an external hard drive with password protection in an enclosed locker. We replicated the procedure when examining the left breast. While the participants were dressing up after the examination, the SS documented additional pertinent notes about the observed breasts such as density of the tissue, presence of fibrotic tissue, evidence of mass or tissue with tumor-related characteristics. Lastly, the participants responded a survey to evaluate their comfort using the device and the potential for adopting the PLB as a routinely screening tool between mammography annual scheduling. The SS placed the results in the corresponding previously coded envelop, sealed it, and stored it in a locked file cabinet. The Principal Investigator (PI) stated the data analysis by opening the matched coded envelopes (radiologist certified mammography results and PLB results as reported by the SS) and creating a data base using Microsoft Office Excel application. Once verified by another colleague, we sent the results to our co-PI’s for statistical analyses.

### Sample Size

For this study, we used the sample size estimator from Epidat v 4.2: Estimation for diagnostic test. We use the sensitivity and sensitivity reported by Bundred and Levack (1986)) and sensitivity and specificity for mammography examinations based on Breast Cancer Research Consortium, 2007-2013 (2017). With a power of 80% and an alpha of 95%, the estimated sample size was 296 examinations with 296 mammograms.

### Statistical Analysis

Each breast was considered an independent event; a total of 340 events were used in the evaluation of the PLB test performance. The mean standard error age at enrollment was 53.3 with a range of 40-77 years. Descriptive statistics were performed using Stata v 16 for the demographic variable of age and questions related to the acceptability of screening tests for early detection of breast cancer, including acceptability of the device under study (PLB). Sensitivity, specificity, positive predictive value, negative predictive value, Youden Index, prevalence and likelihood ratio was assessed using Epidat v. 3.1. The adjusted results of the screening test were evaluated using the imperfect standard analysis, using mammography specificity and sensitivity (BCC, 2020).

## RESULTS

At Table 1, we present the adjusted values by the imperfect reference standard sensitivity for the PLB device 89.6% (95%CI, 84.8-94.3), and specificity of 96.4% (96%CI, 81.3-115.9). Positive predictive value was of 98.8% (95%CI 92.8, 104.4) and negative predictive value 74.4 (95%CI 62.7, 86.1), youden index was 0.9 (95%CI 0.7, 1.1), after adjustment by the imperfect reference standard (mammography). Kappa statistic was 0.54, pre-test probability was of 54.7 while the posttest probability was of 98.2%.

**Table 1.**
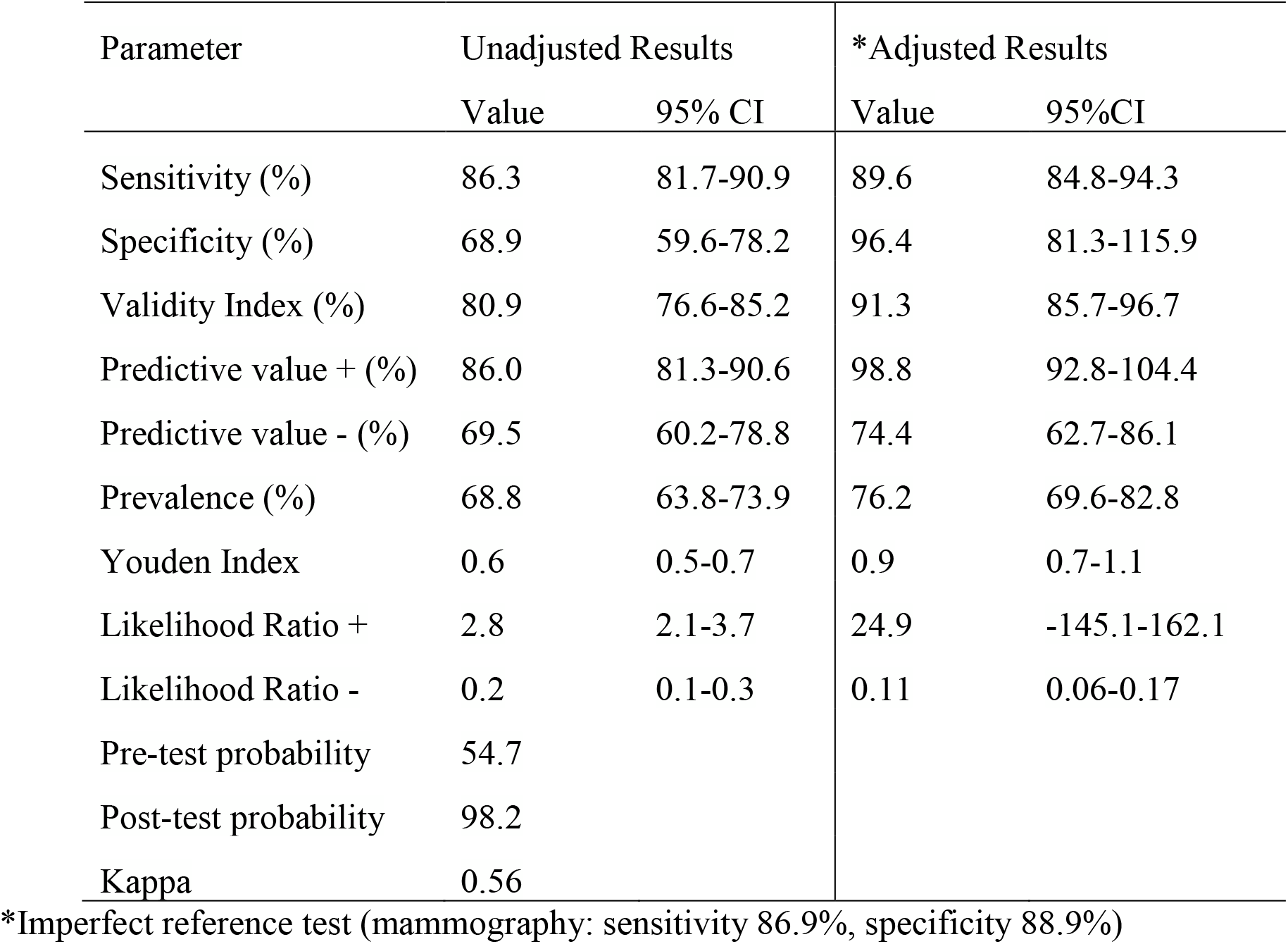
Evaluation of PLBTest Performance with an Imperfect Reference Standard: Mammography.

The data of Table 2 show the acceptability-related questions for breast cancer screening tools by the participants. Participants were women with a mean age of 53.3 SD 9.9, age range of the participants was from 40 years to 77 years. Among the questions related to the acceptability of screening test, 97% (n=164) of the participants reported being comfortable taking a screening test. All participants (100%) who completed the survey (n=169), answered that the PLB device was easy to use a to visualize the breast. The question related to discomfort caused by the device, 100% (n=169) reported none discomfort caused by the device. The item related to reasons to avoid mammography’s, the 70.4% (n=119), reported they were afraid they might hurt, followed by anxiety 20.1% (n=34), and 9.5% (n=16), reported they did not like mammography’s as a screening tool.

**Table 2.** Acceptability of breast cancer screening tools (n= 169)

|  | n (%) |
| --- | --- |
| Age |  |
| Mean, (SD) | 53.3 (9.9) |
| Range | 40-77 |
| I feel comfortable taking screening tests? | 164 (97.0) |
| Is the PLB device easy to use and to visualize my breasts? | 169 (100.0) |
| The discomfort felt by the PLB device was: |  |
| None | 169 (100.0) |
| Reasons to avoid a mammography |  |
| I don't like it | 16 (9.5) |
| Afraid might hurt | 119 (70.4) |
| Anxiety | 34 (20.1) |

PLB: Pink Luminous Breast

Based on the limitation of the mammography for detection of breast abnormalities, Table 3 presents the percentage of positive dense findings with mammography and PLB device, from a total of 340 events, the mammography detected 248 dense breast (72.9%) and PLB device detected 266 dense breasts (78.2%).

**Table 3:** Percentage of positive dense findings of mammography and PLB (n=340)

|  | Positive n (%) |
| --- | --- |
| **Mammography** | 248 (72.9) |
| **PLB Device** | 266 (78.2) |

PLB: Pink Luminous Breast

## DISCUSSION

Breast cancer still leading the cancer cause of death among women and with an increasing trend of prevalence and mortality rate globally (Azamjah, Soltan-Zadeh, & Zayeri, 2019). Currently, screening with mammography is the most effective method to detect early stage of the disease and decrease mortality (Coleman, 2017). However due to its limitations there exist the need for other breast screening tools that not only exhibit high sensitivity and specificity but also affordability, acceptance by users, and easy to use. Issuing of new safe, accurate, and cost-effective tools for breast screening are essential for appropriate decision making. Accessibility to women to self-monitor her breast with a sensitive tool, will contribute to create awareness in women, increasing the probabilities of visit their physicians and acting as an adjunct tool with the mammography, look up for improving prognosis by promoting earlier interventions. Breast screening with the PLB device is non-invasive, with short time of implementation, and with high probability of its use due to the lack or minimum discomfort during examination. We estimated the screening accuracy of PLB in detecting breast abnormal or suspicious areas in the breast to be 81% with a sensitivity of 89.6%, a specificity of 96.4%, a positive predictive value of 98.8%, and a negative predictive value of 74.4%. Comparable values to those for a mammography (Zeeshan, et al., 2018) but with the advantage to be portable, rapid, and painless. Importantly, the PLB allowed the breast classification of density in 78.2% of the studied breast vs a 72.9 the mammography. As we used the intermediate light setting of the PLB for variable control purposes, it will be of great importance to further explore if at a higher light setting, the PLB can be able to detect those lesions missed by the mammography alone.

A limitation of PLB device as well as the mammography is the inability to distinguish benign from malignant breast findings, lacking the outcomes of the pathological examination. In a future we expect to overcome these limitations by using a prospective approach following the final diagnosis with both the mammography and the PLB alone and together. Overall however, to our knowledge there are limited studies addressing the long-term survival of women whose breast cancers were detected with supplemental imaging modalities (Khan, 2021). On the other hand, the strategy used to study the capability of the PLB device on detecting suspected areas was a double-blind approach, a strategy used commonly among clinical trials. All strategies from protecting information, not sharing results with participant, the use of unidentified sealed envelopes and not allowing participants to comment their results were some of the strategies that were implemented to obtain more objectives results.

## CONCLUSSIONS

PLB device has a high sensitivity (89.6%) and specificity (96.4%) for detecting breast abnormalities comparable to the adjusted mammography values: 86.3% and 68.9% respectively. Considering this study significant outcomes, we validated the PLB with the potential to be recommended to be used routinely for breast screening at non-clinical settings. The PLB device provides a rapid, non-invasive, portable, and easy-to-use tool for breast screening that can complement the home-based BSE technique or the CBE. Importantly, the PLB can be conveniently used for screening breast with implants, for which we plan to study this population in a near future. The use of this device is not intended to replace the mammography as the gold standard for breast screening but rather to be used an adjunct or complement tool for earlier detection strategies. At this stage, the main objective for the positive validation outcomes of the PLB is to provide an accessible and painless breast cancer screening tool along to promote awareness about the importance of frequent screening for early detection of breast abnormalities to decrease this health disparity.

## Data Availability

The numbers for mortality and survival ratesused in this manuscript were retrieved from the American Cancer Society, Center for Disease Control and guidelines, reports and bulletins from relevant Societies such as ACS, AAFP, USPSTF, ACOG, NCCN

https://www.cancer.org/cancer/breast-cancer/screening-tests-and-early-detection.html

https://www.cancer.org/content/dam/cancer-org/research/cancer-facts-and-statistics/annual-cancer-facts-and-figures/2021/cancer-facts-and-figures-2021.pdf

https://gis.cdc.gov/Cancer/USCS/DataViz.html

https://www.uspreventiveservicestaskforce.org/uspstf/recommendation/breast-cancer-screening

https://www.aafp.org/afp/2020/0201/p184.html

https://www.acog.org/clinical/clinical-guidance/practice-bulletin/articles/2017/07/breast-cancer-risk-assessment-and-screening-in-average-risk-women

https://www.nccn.org/evidenceblocks/Default.aspx

https://apps.who.int/iris/handle/10665/254500

## ACKNOWLEDEGEMENTS

Our most sincere gratitude to the University of Puerto Rico ay Ponce NIH-NIGMS UPR-PRISE Program undergraduate students Marinel Ocasio, Emmanuel Leon, and Maria del Mar Mendez your support at editing this manuscript.

